# Sleep Hygiene Index: Dimensionality, internal consistency, and nomological validity among Colombian medical students

**DOI:** 10.1101/2024.02.28.24303514

**Authors:** Adalberto Campo-Arias, John Carlos Pedrozo-Pupo, Carmen Cecilia Caballero-Domínguez

## Abstract

**Background and purpose:** A new version of the Sleep Hygiene Index (SHI-10) has recently been introduced, and the psychometric performance in other populations is unknown. This study aimed to determine the dimensionality, internal consistency, and nomological validity of the SHI-10 among medical students at a Colombian university.

**Methods:** A psychometric study was designed to determine indicators of validity (construct and nomological) and reliability (internal consistency) in which 309 medical students between 18 and 39 years (M=20.83, SD=2.68), and 54.69% were female. Construct validity (dimensionality) was tested through confirmatory factor analysis, internal consistency with Cronbach’s alpha and McDonald’s omega coefficients, and nomological validity through correlations with the Athens Insomnia Scale, Epworth Somnolence Scale, Generalized Anxiety Disorder (GAD)-7) and Patient Health Questionnaire (PHQ-9).

**Results:** The four-dimensional structure of the SHI-10 showed adequate indicators of goodness of fit (Satorra-Bentler’s chi-square of 43.30 [df of 29, p=.04], chi-square/df of 1.49, RMSEA of .04 [90%CI .01-.06], CFI of .97, TLI of .96 and SRMR .04). The four dimensions of the SHI-10 showed values less than .70 and limited nomological validity (most Pearson correlations were less than .30).

**Conclusions:** The SHI-10 shows a four-dimensional structure of SHI-10; however, the four dimensions of the SHI-10 present low internal consistency and limited nomological validity. More studies are needed to show the psychometric performance of the SHI-10.

## INTRODUCTION

Sleep hygiene refers to the activities of daily living that promote good sleep quality and full daytime alertness (De Biase et al., 2014). In college students, poor nighttime sleep is associated with daytime sleepiness, feelings of tiredness, changes in mood, impairment in global functioning, and other adverse health effects (Anwer et al., 2019; Irish et al., 2015; Kang & Chen, 2009; Kaur & Singh, 2017; Manzar et al., 2020).

### SLEEP HYGIENE INDEX

One of the instruments with the best psychometric indicators to quantify habits that affect sleep is the thirteen-item Sleep Hygiene Index (SHI-13). In the presentation article, the SHI-13 showed acceptable internal consistency, Cronbach’s alpha of .66, high test-retest stability, *r* of .71 (p<.01), and acceptable nomological validity, with a modest high correlation with the Epworth Sleepiness Scale (ESS), *r* of .24 (p<.01) and adequate correlation with the Pittsburgh Sleep Quality Index, *r* of .48 (Mastin et al., 2006). These characteristics have been corroborated in other studies with the participation of university students, particularly internal consistency values, Cronbach’s alpha, or McDonald’s omega, less than .70 (Ali et al., 2021; Kaur & Singh, 2017; Seun-Fadipe et al., 2018; Tang et al., 2021). Generally, internal consistency values greater than .70 are recommended (Campo-Arias & Oviedo, 2008; Kezsei et al., 2010).

Another limitation observed for the SHI-13 has been the poor reproducibility of dimensionality. Mastin et al. (2006) did not explore the dimensionality of the SHI-13, and between three and six dimensions have been documented in different populations (Ali et al., 2021; Anwer et al., 2019; Seun-Fadipe et al., 2018; Tang et al., 2018; 2021). Theoretically, these disparities in the dimensions of the SHI-13 undermine the overall validity of the instrument and limit its usefulness in clinical and epidemiological studies (Streiner & Norman, 2008).

Due to the limitations noted above, Prados et al. (2021) refined the SHI-13. After eliminating items 1, 4, and 6 due to low factor loadings, they observed that a 10-item version (SHI-10) showed better performance in 548 Spanish university students: The Exploratory factor analysis of the SHI-10 using the principal components extraction method showed a solution of four factors that explained 65.58% of the total variance (factor 1 28.11%, factor 15.00%, factor 3 11.63% and factor 4 10.84%). The factors were: Factor 1 (sleep-disrupting behaviors, items 5, 7 and 9, with McDonald’s omega of 0.75), factor 2 (cognitive activation, items 8, 12 and 13; with McDonald’s omega of 0.75), 78); factor 3 (bedroom comfort, items 10 and 11, with McDonald’s omega of 0.88) and factor 4 (sleep/wake time, items 2 and 3, with McDonald’s omega of 0.83). Likewise, nomological validity was limited because only factor 2 (cognitive activation) showed significant correlations, more excellent than .30, with the Depression, Anxiety, and Stress Scale (DASS-21) and the Pittsburgh Sleep Quality Index (PSQI). Consequently, the dimensions of the HSI-10 showed better internal consistency than that observed in other studies for the global SHI-13 (Ali et al., 2021; Kaur & Singh, 2017; Mastin et al., 2006; Seun-Fadipe et al., 2018; Tang et al., 2021).

Likewise, the four-dimensional structure of SHI-10 is similar to that reported by Anwer et al. (2019) for the SHI-13. However, factors 3 (bedroom comfort) and 4 (sleep/wake time) can be problematic because they only contain two items. The reliability of a factor is more likely guaranteed if this factor has at least three items (Campo-Arias et al., 2012; Gorsuch, 1997; Streiner, 1994).

### THE PRESENT STUDY

The psychometric performance of health measurement instruments is highly variable according to the population’s characteristics, which usually undermines the validity and reliability of the measurements made (Streiner & Norman, 2008). In the present study, to expand the knowledge of the psychometric performance of the SHI-10, a confirmatory factor analysis (CFA) is carried out in Colombian medical students using the maximum likelihood extraction method; this confirmation was omitted in the group of Spanish students (Prados et al., 2021). Furthermore, the extraction by the maximum likelihood method used in the CFA is indicated for the extraction of factors because the principal component method is indicated for the reduction of the number of unrelated variables as the items of a given item are expected to be a scale that measures the same construct (Gorsuch, 1997; Norman & Streiner, 2008; Streiner, 1994). Likewise, two coefficients will be calculated for internal consistency following the most recent recommendations in psychometrics (American Educational Research Association et al., 2014; International Test Commission, 2017).

Finally, to test nomological validity, the correlation of the scores of the HSI-10 dimensions with the scores on the Athens Insomnia Scale [AIS] (Soldatos et al., 2000), the Epworth Sleepiness Scale [ESS] (Johns, 1994) will be explored, the Generalized Anxiety Disorder [GAD-7 for anxiety] (Spitzer et al., 2006) and the Patient Health Questionnaire [PHQ-9 for depression] (Kroenke et al., 2001). These measurements were taken because it is documented that sleep problems are significantly correlated with scores for anxiety or depression symptoms (Ghrouz et al., 2019; Vanderlind et al., 2014).

### PRACTICAL CONSIDERATIONS

Sleep hygiene is essential in preventing and managing sleep disorders (American Academy of Sleep Medicine, 2023; De Biase et al., 2014). More than 50% of college students report poor sleep quality around the world (Abdulghani et al., 2012; Bahammam et al., 2012; Brick et al., 2010; Feng et al., 2005; Lu et al., 2011; Manzar et al., 2019; Preišegolavičiūtė et al., 2010) due to inadequate sleep hygiene associated with academic demands, especially during exam times and rotating shifts like medical students (Ahrberg et al., 2012; Hershner & Chervin, 2014). The deterioration in the sleep pattern negatively affects cognitive processes (Ahrberg et al., 2012; Giri et al., 2013; Ratcliff & Van Dongen, 2009) and, consequently, academic performance and achievements (Menon et al., 2015; Short et al., 2013; Vanderlind et al., 2014).

### AIM

Determine the dimensionality, internal consistency, and nomological validity of the SHI-10 among medical students at a university in Santa Marta, Colombia.

## METHODS

### Study design

A psychometric study was implemented to determine indicators of validity (construct and nomological) and reliability (internal consistency).

### Participants

The authors invited adult students to participate voluntarily. From the first to the tenth semester, three hundred nine medical students agreed to participate in the study. This number of participants was adequate for evaluating a 10-item scale at a ratio superior to 20 participants for each SHI item (Campo-Arias et al., 2012). The ages of students were observed between 18 and 39 years (20.83±2.68). The most significant participants were students under 20, female students, low-income students, and from urban areas. More details of the participants’ characteristics are presented in Table 1.

**Table 1.** Demographic characteristics of participating students.

| Variable | Frequency | % |
| --- | --- | --- |
| *Age* |  |  |
| Between 18 and 19 | 166 | 53.72 |
| 20 or more | 143 | 46.28 |
| *Gender* |  |  |
| Female | 169 | 54.69 |
| Male | 140 | 45.31 |
| *Income* |  |  |
| Low | 194 | 62.78 |
| High | 115 | 37.22 |
| *Origin residence* |  |  |
| Urban | 266 | 86.08 |
| Rural | 43 | 13.92 |
| *Semester* |  |  |
| First | 29 | 9.51 |
| Second | 23 | 7.43 |
| Third | 19 | 6.14 |
| Fourth | 32 | 10.34 |
| Fifth | 24 | 7.76 |
| Sixth | 53 | 17.14 |
| Seventh | 46 | 14.88 |
| Eighth | 32 | 10.35 |
| Nineth | 22 | 7.11 |
| Tenth | 27 | 8.74 |

### Instruments

#### SHI-10

The SHI-10 comprises ten items that explore behaviors that can affect the quantity and quality of sleep. These items contribute to four factors: Factor 1 (sleep-disrupting behaviors, items 3, 4, and 6), factor 2 (cognitive activation, items 5, 9, and 10), factor 3 (bedroom comfort, items 7 and 8), and factor 4 (sleep/wake time, items 1 and 2 (Prados et al., 2021).

A version with five response options was applied, from never to always, and is rated from zero to four with total scores between 0 and 40; the higher the score, the worse the sleep hygiene (Prados et al., 2021).

#### AIS

The AIS is an inventory quantifying sleep difficulties related to insomnia over the past month. The AIS consists of eight items with four response options, scored from zero (meaning it is not a problem) to three (most acute difficulty sleeping), with total scores between 0 and 24; the higher the score, the more insomnia, and associated symptoms (Soldatos et al., 2000). The AIS has shown high internal consistency in previous Colombian research (Pedrozo-Pupo et al., 2022). In the present study, the AIS showed Cronbach’s alpha of .80.

#### ESS

The ESS is an eight-item instrument that quantifies the probability of falling asleep in eight daily situations. The EES provides four response options scored between zero and three, with total scores between 0 and 24 (Johns, 1994). This instrument has presented acceptable dimensionality and internal consistency in Colombian studies (Pedrozo-Pupo et al., 2020). The ESS showed Cronbach’s alpha of .83 in this present group of students.

#### GAD-7

The GAD-7 is a seven-item scale to assess anxiety symptoms during the most recent two weeks. This instrument presents four response options from “none of the days” to “almost every day” that are scored from zero to three, the higher the anxiety score (Spitzer et al., 2006). The GAD-7 presented high internal consistency in a previous Colombian study (Monterrosa-Blanco et al., 2021). In the present sample, the GAD-7 showed Cronbach’s alpha of .91.

#### PHQ-9

The PHQ-9 explores symptoms of a major depressive episode in the past 15 days. The PHQ-9 is made up of nine questions with four response options from “not at all” to “almost every day” that are rated from zero to three; the higher the score, the greater the depression (Kroenke et al., 2001). This instrument has shown adequate validity and reliability indicators in studies in Colombia (Cassiani-Miranda & Scoppetta, 2018). Cronbach’s alpha was .88 in the present research.

### Procedures

Students completed an online questionnaire in the classroom on a group application after a research assistant explained the study’s objectives and that participation was voluntary. This information was completed in the second half of 2022.

### Statistical analysis

#### Dimensionality

The dimensionality of the SHI-10 was tested using CFA, with the maximum likelihood extraction method and Promax rotation. The Promax rotation is the most indicated when the theoretical assumption is made that the factors or dimensions of an instrument have a high correlation between them (Streiner, 1993). The loadings were observed, and the goodness-of-fit indicators were calculated: Satorra-Bentler’s chi-square and normalized chi-square, Root Mean Square Error of Approximation (RMSEA) with a 90% confidence interval (90%CI), Comparative Fit Index (CFI), the Tucker-Lewis index (TLI) and the Standardized Mean Square Residual (SRMR).

It is desirable to observe a value of normalized chi-square less than 3.00 (Hair et al., 2006). RMSEA is acceptable if it is less than 0.06, the CFI and TLI should be greater than 0.90, and the SRMR is expected to be less than 0.05 (Hu & Bentler, 1999).

#### Internal consistency

Internal consistency was determined and calculated with Cronbach’s alpha (1951) and McDonald’s omega (1970) coefficients. McDonald’s omega is a better indicator of internal consistency when the tau-equivalence principle necessary for the calculation of Cronbach’s alpha is not met; that is, the factor loadings of the items are similar (Hayes & Coutts, 2020; Trizano-Hermosilla & Alvarado, 2016).

#### Nomological validity

Nomological or hypothesis validity was estimated with Pearson’s (1909) correlation of the point of each dimension of the SHI and the total scores on the AIS, the ESS, GAD-7, and PHQ-9. Those with a r value equal to or greater than .30, with a probability value lower than .01, were accepted as significant correlations (Streiner & Norman, 2008).

### Ethical considerations

This project was approved by an institutional ethics committee of the Universidad del Magdalena at Santa Marta, Colombia, according to minutes 005 of the ordinary virtual session of June 9, 2022. The students signed an online informed consent form, and measurement scales were used for free, in line with national and international standards for research involving humans (World Medical Association, 2018).

## RESULTS

### Dimensionality

The four-dimensional structure of SHI-10 was corroborated. Satorra-Bentler’s chi-squared of 43.30 (df of 29, p=.04), normalized chi-squared of 1.49, CFI of 97, TLI of .96, and SRMR .04. The loadings for the SHI-10 items were observed between .40 and .81. All loadings are presented in Table 2.

**Table 2.**
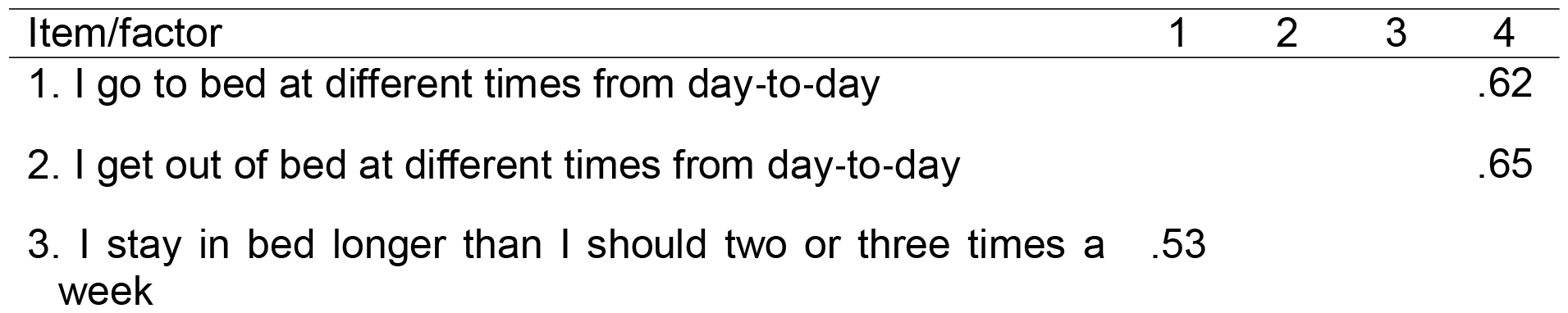
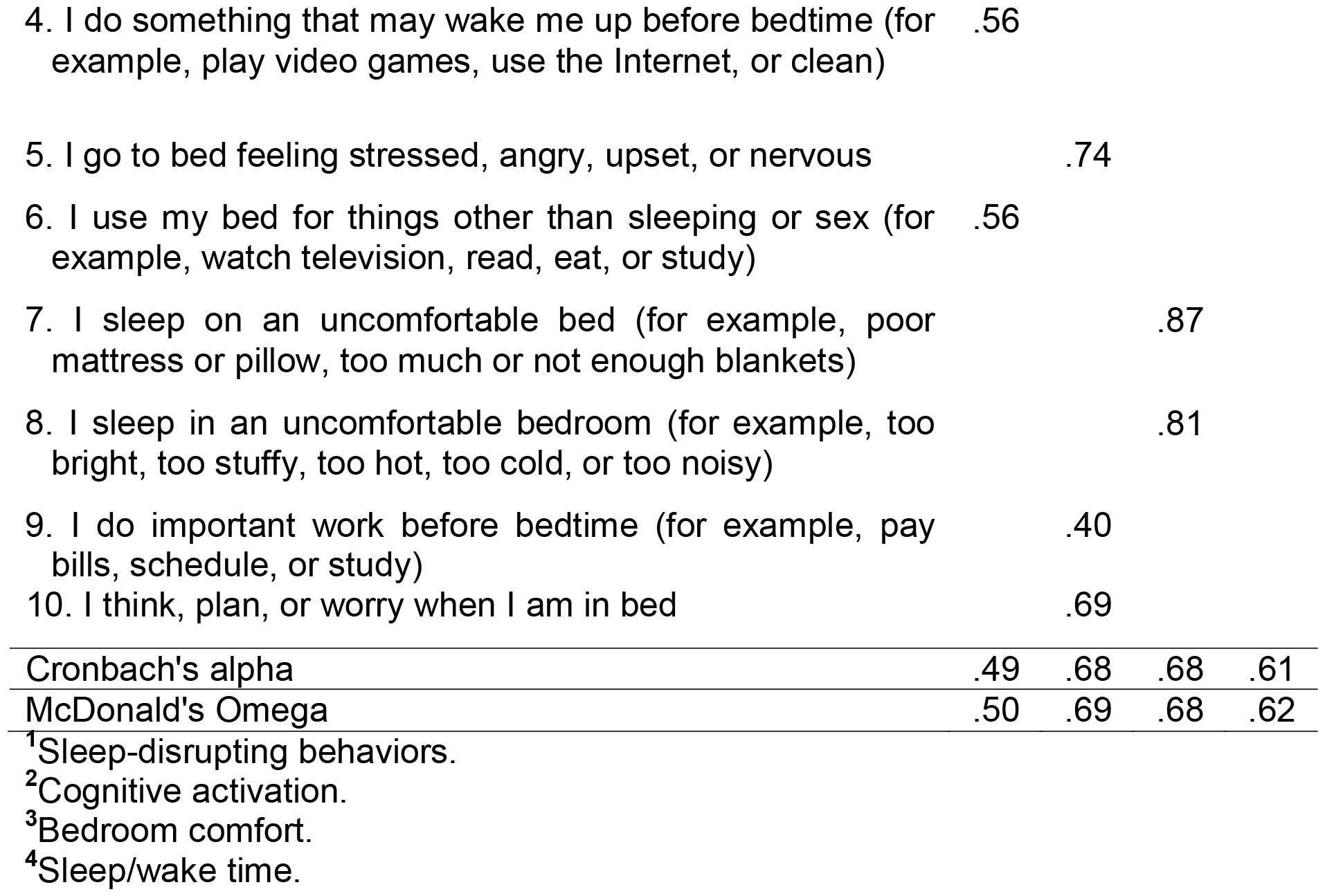
Confirmatory factor analysis using the maximum likelihood method and promax rotation for the four-dimensional solution.

### Internal consistency

The internal consistencies of the dimensions of the four dimensions of the SHI-10 were observed between .49 (factor 1, sleep-disrupting behaviors) and .68 (factors 2 and 3, cognitive activation and bedroom comfort) for Cronbach’s alpha and .50 (factor 1, sleep-disrupting behaviors) and .69 (factor 2, cognitive activation) for McDonald’s omega. See the coefficients at the bottom of Table 2. Overall, the HSI-10 items showed Cronbach’s alpha of .72 and McDonald’s omega of .73.

### Nomological validity

The scores in the AIS were between 0 and 21 (8.88±3.97), in the ESS between 0 and 24 (11.63±4.92), in the GAD-7 between 0 and 21 (7, 92±5.36), in the PHQ-9 between 0 and 27 (8.94±5.95) and in the HSI-10 between 0 and 38 (24.04±5.55). These scores in factor 1 were observed between 1 and 12 (7.61±2.65), in factor 2 between 2 and 12 (8.37±2.20), in factor 3 between 0 and 8 (2 .39±2.04), and factor 4 between 1 and 8 (5.70±1.63).

The correlations of the HSI dimensions with the scores in AIS, ESS, GAD-7, and PHQ-9 between .09 and .56. Factor 1 (sleep-disrupting behaviors) and factor 3 (bedroom comfort) showed poor correlations with all scales, and factor 2 (cognitive activation) presented statistically significant coefficients with all scales. See the values in Table 3.

**Table 3.** Correlations between factors and scores for insomnia (AIS), somnolence (ESS), anxiety (GAD-7), and depression (PHQ-9).

SLEEP HYGIENE INDEX-10
| Factor/variable | AIS | ESS | GAD-7 | PHQ-9 |
| --- | --- | --- | --- | --- |
| 1. Sleep-disrupting behaviors | .24 | .19 | .20 | .24 |
| 2. Cognitive activation | .54* | .43* | .56* | .51* |
| 3. Bedroom comfort | .26 | .09 | .19 | .21 |
| 4. Sleep/wake time | .37* | .17 | .27 | .34* |

\*The correlation is significant at the .01 level (two-tailed).

The SHI-10 total scores showed statistically significant Pearson correlations (p<.01) with the total scores for insomnia (*r* =.53), excessive sleepiness (*r* =.34), anxiety (*r* =.46), and depression (*r* =.48).

## DISCUSSION

The current study shows that the four-dimensional structure of the SHI-10 is unlikely among medical students from Santa Marta, Colombia. The four dimensions of the SHI-10 show low consistency and limited nomological validity compared to scores for anxiety, depression, insomnia, and somnolence.

The present study’s findings are sufficiently similar to what was reported in the introductory article of the SHI-10 concerning the four-dimensionality of the instrument and the low correlations of the dimension scores with external measurements. However, internal consistencies were found in the data analyzed, measured with two coefficients below the minimum expected values (Prados et al., 2021).

Ideally, a valid instrument is expected, for example, in dimensionality and nomological and reliable validity, with high internal consistency (American Educational Research Association et al., 2014; International Test Commission, 2017; Norman & Streiner, 2008). An instrument with limited validity and poor reliability has limited usefulness in the measurements (Streiner & Norman, 2008). This point suggests the need to review the construct, the theoretical basis that supports the instrument, or the content or wording of the items (Gorsuch, 1997; Keszei et al., 2010), for example, considering the possibility of designing a unidimensional scale or dimensions with three or more items; consequently, having a four-dimensional structure for the 10-item version of the SHI is complex (Campo-Arias et al., 2012; Gorsuch, 1997; Streiner, 1993). The discrepancy between the validity indicators and the reliability of a measurement scale constitutes a challenge for professionals in clinical and epidemiological studies, mainly when there is a lack of a reference criterion to test the performance of an instrument (Rush et al., 2009; Norman & Streiner, 2008).

### PRACTICAL CONSIDERATIONS

Health professionals must remember the need to corroborate the validity and reliability of measurement instruments. The findings show that the SHI’s validity and reliability are questionable in Colombian medical students. The ‘validation’ process of health measurement scales requires a permanent review and refinement to make the necessary adjustments so that the validity and reliability indicators are within the recommended parameters (Campo-Arias & Pineda-Roa, 2022). Likewise, it must be kept in mind that the validity and reliability indicators of the scales may present significant divergences between different population groups (Keszei et al., 2010; Norman & Streiner, 2008). Consequently, it may be more appropriate to report ‘psychometric performance’ than ‘psychometric properties’ or ‘validation’ of a measurement scale, given that it refers more to the response pattern of a population than to the intrinsic characteristics of the instrument (Campo-Arias & Oviedo, 2008; Campo-Arias & Pineda-Roa, 2022; Keszei et al., 2010; Norman & Streiner, 2008). Psychometric theory is the basis for developing clinical mental health research assessment instruments. However, the psychometric model shows limitations for concepts more related to the clinical one due to the high heterogeneity of the populations (Fava et al., 2004).

### STUDY’S STRENGTHS AND LIMITATIONS

This study is advanced by performing a CFA, calculating two indicators of internal consistency (Cronbach’s alpha and McDonald’s omega). The SHI-10 introductory paper omitted this approach (Prados et al., 2021). An attempt was made to test the performance of the SHI-10 in people from a different cultural context. Cultural aspects can significantly affect the response pattern in a measurement instrument (American Educational Research Association et al., 2014; International Test Commission, 2017). Additionally, it extensively tested the nomological validity of the SHI-10 dimensions or factors by looking at correlations with scores for anxiety, depression, insomnia, and sleepiness. However, this research has the same limitation as psychometric studies that only apply to the participating sample without being able to generalize to other populations, although these populations, in theory, are considered similar (Keszei et al., 2010; Norman & Streiner, 2008). It is necessary to continue exploring the psychometric performance of the SHI-10 in different populations or cultural contexts (Campo-Arias & Pineda-Roa, 2022).

## CONCLUSIONS

The four-dimensional structure of the SHI-10 took much work to accept among Colombian medical students. The internal consistencies of the four factors were observed below recommended values, and nomological validity was only demonstrated for factor 2 (cognitive activation). Testing other structures that show dimensions with high internal consistency and acceptable nomological validity is necessary.

## Data Availability

The corresponding author can request data supporting this research with reasonable justification.

## REFERENCES

Abdulghani, H. M., Alrowais, N. A., Bin-Saad, N. S., Al-Subaie, N. M., Haji, A. M., & Alhaqwi, A. I. (2012). Sleep disorder among medical students: Relationship to their academic performance. Medical Teacher, 34(Supl.1), S37–S41. 10.3109/0142159X.2012.656749

Ahrberg, K., Dresler, M., Niedermaier, S., Steiger, A., & Genzel, L. (2012). The interaction between sleep quality and academic performance. Journal of Psychiatric Research, 46(12), 1618–1622. 10.1016/j.jpsychires.2012.09.008

Ali, R., Zolezzi, M., & Awaisu, A. (2021). The Arabic version of the sleep hygiene index: Linguistic validation and cultural adaptation among university students in Qatar. Qatar Medical Journal, 2021(2), 26. 10.5339/qmj.2021.26

American Academy of Sleep Medicine. (2023). International Classification of Sleep Disorders, 3rd edition. American Academy of Sleep Medicine.

American Educational Research Association, American Psychological Association, & National Council on Measurement in Education. (2014). Standards for educational and psychological testing. American Educational Research Association.

Anwer, S., Alghadir, A., Manzar, M. D., Noohu, M. M., Salahuddin, M., & Li, H. (2019). Psychometric analysis of the sleep hygiene index and correlation with stress and anxiety among Saudi University students. Nature and Science of Sleep, 11, 325–332. 10.2147/NSS.S222440

BaHammam, A. S., Alaseem, A. M., Alzakri, A. A., Almeneessier, A. S., & Sharif, M. M. (2012). The relationship between sleep and wake habits and academic performance in medical students: A cross-sectional study. BMC Medical Education, 12(1), 60. 10.1186/1472-6920-12-61

Brick, C. A., Seely, D. L., & Palermo, T. M. (2010). Association between sleep hygiene and sleep quality in medical students. Behavioral Sleep Medicine, 8(2), 113–121. 10.1080/15402001003622925

Campo-Arias, A., & Oviedo, H. C. (2008). Propiedades psicométricas de una escala: la consistencia interna [Psychometric properties of a scale: Internal consistency]. Revista de Salud Pública, 10(5), 831–839.

Campo-Arias, A., & Pineda-Roa, C. A. (2022). Instrument validation is a necessary, comprehensive, and permanent process. Alpha Psychiatry, 23(2), 89–90. 10.5152/alphapsychiatry.2022.21811

Campo-Arias, A., Herazo, E., & Oviedo, H. C. (2012). Análisis de factores: fundamentos para la evaluación de instrumentos de medición en salud mental [Factor analysis: Principles to evaluate measurement tools for mental health]. Revista Colombiana de Psiquiatría, 41(3), 659–671. 10.1016/S0034-7450(14)60036-6

Cassiani-Miranda, C. A., & Scoppetta, O. (2018). Factorial structure of the Patient Health Questionnaire-9 as a depression screening instrument for university students in Cartagena, Colombia. Psychiatry Research, 269, 425–429. 10.1016/j.psychres.2018.08.071

Cronbach, J. (1951). Coefficient alpha and the internal structure of tests. Psychometrika, 16(3), 297–334. 10.1007/bf02310555

De Biase, S., Milioli, G., Grassi, A., Lorenzut, S., Parrino, L., & Gigli, G. L. (2014). Sleep hygiene (pp. 289–295). In S. Garbarino, L. Nobili & G. Costa. Sleepiness and Human Impact Assessment. Springer.

Fava, G. A., Ruini, C., & Rafanelli, C. (2004). Psychometric theory is an obstacle to the progress of clinical research. Psychotherapy and Psychosomatics, 73(3), 145–148. 10.1159/000076451

Feng, G. S., Chen, J. W., & Yang, X. Z. (2005). Study on the status and quality of sleep-related influencing factors in medical college students. Chinese Journal of Epidemiology, 26(5), 328–331.

Ghrouz, A. K., Noohu, M. M., Manzar, M. D., Spence, D. W., BaHammam, A. S., & Pandi-Perumal, S. R. (2019). Physical activity and sleep quality in relation to mental health among college students. Sleep Breath, 23(2), 627–634. 10.1007/s11325-019-01780-z

Giri, P. A., Baviskar, M. P., & Phalke, D. B. (2013). Study of sleep habits and sleep problems among medical students of Pravara Institute of Medical Sciences Loni, Western Maharashtra, India. Annals of Medical and Health Sciences Research, 3(1), 51–54. 10.4103/2141-9248.109488

Gorsuch, R. L. (1997). Exploratory factor analysis: Its role in item analysis. Journal of Personality Assessment, 68(3), 532–560. 10.1207/s15327752jpa6803_5

Hair, J., Black, B., Babin, B., Anderson, R., & Tatham, R. (2006). Multivariate data analysis. 6th edition. Prentice-Hall.

Hayes, A. F., & Coutts, J. J. (2020). Use omega rather than Cronbach’s alpha for estimating reliability. But…. Communication Methods and Measures, 14(1), 1–24. 10.1080/19312458.2020.1718629

Hershner, S. D., & Chervin, R. D. (2014). Causes and consequences of sleepiness among college students. Nature and Science of Sleep, 6, 73–84. 10.2147/NSS.S62907

Hu, L. T., & Bentler, P. M. (1999). Cutoff criteria for fit indexes in covariance structure analysis: Conventional criteria versus new alternatives. Structural Equation Modeling, 6(1), 1–55. 10.1080/10705519909540118

International Test Commission. (2017). ITC guidelines for translating and adapting tests (2nd edition). http://www.InTestCom.org

Irish, L. A., Kline, C. E., Gunn, H. E., Buysse, D. J., & Hall, M. H. (2015). The role of sleep hygiene in promoting public health: A review of empirical evidence. Sleep Medicine Reviews, 22, 23–36. 10.1016/j.smrv.2014.10.001

Johns, M. W. (1994). Sleepiness in different situations measured by the Epworth Sleepiness Scale. Sleep, 17(8), 703–710. 10.1093/sleep/17.8.703

Kang, J. H., & Chen, S. C. (2009). Effects of an irregular bedtime schedule on sleep quality, daytime sleepiness, and fatigue among university students in Taiwan. BMC Public Health, 9(1), 248. 10.1186/1471-2458-9-248

Kaur, G., & Singh, A. (2017). Sleep hygiene, sleep quality and excessive daytime sleepiness among Indian college students. Journal of Sleep Medicine & Disorders, 4(1), 1076.

Keszei, A. P., Novak, M., & Streiner, D. L. (2010). Introduction to health measurement scales. Journal of Psychosomatic Research, 68(4), 319–323. 10.1016/j.jpsychores.2010.01.006

Kroenke, K., Spitzer, R. L., & Williams, J. B. (2001). The PHQ-9: validity of a brief depression severity measure. Journal of General Internal Medicine, 16(9), 606–613. 10.1046/j.1525-1497.2001.016009606.x

Lu, J., Fang, G. E., Shen, S. J., Wang, Y., & Sun, Q. (2011). A questionnaire survey on sleeping in class phenomenon among Chinese medical undergraduates. Medical Teacher, 33(6), 508.

Manzar MD, Bekele BB, Noohu MM, Salahuddin, C., Albougami, A., Spence, D. W., Pandi-Perumal, S. R., & Bahammam, A. S. (2019). Prevalence of poor sleep quality in the Ethiopian population: A systematic review and meta-analysis. Sleep Breathing, 10, 709–716. 10.1007/s11325-019-01871-x

Manzar, M. D., Noohu, M. M., Salahuddin, M., Nureye, D., Albougami, A., Spence, D. W., Pandi-Perumal, S. R., & Bahammam, A. S. (2020). Insomnia symptoms and their association with anxiety and poor sleep hygiene practices among Ethiopian university students. Nature and Science of Sleep, 12, 575–582. 10.2147/NSS.S246994

Mastin, D. F., Bryson, J., & Corwyn, R. (2006). Assessment of sleep hygiene using the Sleep Hygiene Index. Journal of Behavioral Medicine, 29(3), 223–227. 10.1007/s10865-006-9047-6

McDonald, R. P. (1970). The theoretical foundations of principal factor analysis, canonical factor analysis, and alpha factor analysis. British Journal of Mathematical and Statistical Psychology, 23(1), 1–21. 10.1111/j.2044-8317.1970.tb00432.x

Menon, B., Karishma, H. P., & Mamatha, I. V. (2015). Sleep quality and health complaints among nursing students. Annals of Indian Academy of Neurology, 18(3), 363–364. 10.4103/0972-2327.157252

Monterrosa-Blanco, A., Cassiani-Miranda, C. A., Scoppetta, O., & Monterrosa-Castro, A. (2021). Generalized anxiety disorder scale (GAD-7) has adequate psychometric properties in Colombian general practitioners during COVID-19 pandemic. General Hospital Psychiatry, 70, 147–148. 10.1016/j.genhosppsych.2021.03.013

Norman, G. R., & Streiner, D. L. (2008). Biostatistics: The bare essentials. D. C. Becker Inc.

Pearson, K. (1909). Determination of the coefficient of correlation. Science, 30(757), 23–25. 10.1126/science.30.757.23

Pedrozo-Pupo, J. C., Caballero-Domínguez, C. C., & Campo-Arias, A. (2022). Prevalence and variables associated with insomnia among COVID-19 survivors in Colombia. Acta Bio Medica, 93(1), e2022019. 10.23750/abm.v93i1.12132

Pedrozo-Pupo, J. C., Córdoba, A. P., & Campo-Arias, A. (2020). Estructura factorial y consistencia interna de la escala de somnolencia de Epworth [Factor structure and internal consistency of the Epworth Sleepiness Scale]. Revista de la Facultad de Medicina, 68(2), 183–187. 10.15446/revfacmed.v68n2.73025

Prados, G., Chouchou, F., Carrión-Pantoja, S., Fernández-Puerta, L., & Pérez-Mármol, J. M. (2021). Psychometric properties of the Spanish version of the Sleep Hygiene Index. Research in Nursing & Health, 44(2), 393–402. 10.1002/nur.22111

Preišegolavičiūtė, E., Leskauskas, D., & Adomaitienė, V. (2010). Associations of quality of sleep with lifestyle factors and profile of studies among Lithuanian students. Medicine, 46(7), 482–489. 10.3390/medicina46070070

Ratcliff, R., & Van Dongen, H. P. (2009). Sleep deprivation affects multiple distinct cognitive processes. Psychonomic Bulletin & Review, 16(4), 742–751. 10.3758/PBR.16.4.742

Rush Jr, A. J., First, M. B., & Blacker, D. (2009). Handbook of psychiatric measures. American Psychiatric Publishing.

Seun-Fadipe, C. T., Aloba, O. O., Oginni, O. A., & Mosaku, K. S. (2018). Sleep hygiene index: Psychometric characteristics and usefulness as a screening tool in a sample of Nigerian undergraduate students. Journal of Clinical Sleep Medicine, 14(8), 1285–1292. 10.5664/jcsm.7256

Short, M. A., Gradisar, M., Lack, L. C., & Wright, H. R. (2013). The impact of sleep on adolescent depressed mood, alertness and academic performance. Journal of Adolescence, 36(6), 1025–1033. 10.1016/j.adolescence.2013.08.007

Soldatos, C., Dikeos, D., & Paparrigopoulos, T. (2000). Athens Insomnia Scale: validation of an instrument based on ICD-10 criteria. Journal of Psychosomatic Research, 48(6), 555–560. 10.1016/S0022-3999(00)00095-7

Spitzer, R. L., Kroenke, K., Williams, J. B., & Löwe, B. (2006). A brief measure for assessing generalized anxiety disorder: The GAD-7. Archives of Internal Medicine, 166(10), 1092–1097. 10.1001/archinte.166.10.1092

Streiner, D. L. (1994). Figuring out factors: the use and misuse of factor analysis. The Canadian Journal of Psychiatry, 39(3), 135–140. 10.1177/070674379403900303

Streiner, D., & Norman, G. (2008). Health measurement scales: A practical guide to their development and use (4th edition). Oxford University Press.

Tang, Z., Li, X., Zhang, Y., Li, X., Zhang, X., Hu, M., & Wang, J. (2021). Psychometric analysis of a Chinese version of the Sleep Hygiene Index in nursing students in China: A cross-sectional study. Sleep Medicine, 81, 253–260. 10.1016/j.sleep.2021.02.050

Trizano-Hermosilla, I., & Alvarado, J. M. (2016). Best alternatives to Cronbach’s alpha reliability in realistic conditions: Congeneric and asymmetrical measurements. Frontiers in Psychology, 7, 769. 10.1016/10.3389/fpsyg.2016.00769

Vanderlind, W. M., Beevers, C. G., Sherman, S. M., Trujillo, L. T., McGeary, J. E., Matthews, M. D., Maddox, W. T., & Schnyer, D. M. (2014). Sleep and sadness: Exploring the relation among sleep, cognitive control, and depressive symptoms in young adults. Sleep Medicine, 15(1), 144–149. 10.1016/j.sleep.2013.10.006

World Medical Association. (2018). WMA Declaration of Helsinki – Ethical principles for medical research involving human subjects. WMA. https://www.wma.net/policies-post/wma-declaration-of-helsinki-ethical-principles-for-medical-research-involving-human-subjects/

